# Nourishing Hearts: The Link Between Food Insecurity and Overall Health Status of Children with Congenital Heart Disease in U.S.

**DOI:** 10.64898/2026.04.03.26350134

**Authors:** Sagar Jani, Hitakshi Modi, Mohit Nadkarni, Charles D Fraser, Davi Freitas Tenorio

## Abstract

**Background:** Children with congenital heart disease (CHD) require specialized care and may face worse outcomes if they experience food insecurity (FI). FI is associated with poor nutrition, hospitalizations, and developmental delays, compounding cardiac risks. Limited research evaluated impact of FI on health status among children with CHD. This study examines socioeconomic factors and the relationship between FI and health status in children with CHD.

**Methods:** 2023 National Survey of Children’s Health (NSCH) data were used to compare rates of FI between children ages ≤ 17 years with and without CHD and to assess overall health status of those with CHD. Descriptive, univariate, and multivariable logistic regression were utilized.

**Results:** Among 53,477 children, 1,233(2%) had CHD. FI was reported in 35% of children with CHD vs. 27% without CHD(p=0.005). After adjustment, children with CHD had higher odds of FI (OR 1.49; 95% CI: 1.05– 2.12). Hispanic ethnicity, residence in Midwest or South, lower household education, and lower poverty index were significantly associated with FI. Households receiving food assistance had higher FI. Living in grandparent household was associated with lower odds of FI. Within the CHD subgroup, 5% reported fair or poor health. Children with CHD experiencing FI had greater odds of fair or poor health than those without FI (OR 3.91, 95% CI 1.70–9.02; p=0.001).

**Conclusions:** Children with CHD face higher odds of FI, which is strongly associated with worse reported health. Addressing socioeconomic vulnerability and FI may improve outcomes and reduce disparities in this high-risk population through targeted screening and intervention strategies nationwide.

## Introduction

Food insecurity (FI), defined by the U.S. Department of Agriculture as the limited or uncertain access to adequate food, affects nearly one in seven U.S. children, disproportionately impacting non-White, low-income, and single-parent households. ^1^ Extensive evidence links FI to adverse pediatric outcomes, including higher rates of hospitalization, poor nutritional intake, obesity, and developmental and behavioral problems.^2–4^ The American Academy of Pediatrics (AAP) emphasizes that FI is a critical child health issue and recommends universal screening using validated tools, such as the Hunger Vital Sign™, to identify and address this modifiable determinant of health.^5^ Pediatricians are uniquely positioned to recognize FI as a health-related social need and to connect families to nutrition and income support programs.^6^

Children with congenital heart disease (CHD) represent a particularly vulnerable group due to their complex metabolic demands, frequent hospitalizations, and heightened nutritional needs.^7,8^ Families caring for children with CHD experience significant financial strain, with more than 89% reporting at least one financial burden, and nearly 15% of caregivers requiring mental health services due to their child’s condition.^9^ The coexistence of FI and CHD may amplify health disparities, as poor nutrition and social stressors can hinder recovery, growth, and neurodevelopment.^10,11^ Despite this, very few studies have investigated the intersection of FI and CHD. This study aims to address this gap by examining the prevalence and predictors of FI among children with CHD in the United States and assessing its association with the overall health status of children with CHD.

## Methods

This cross-sectional study adhered to the Strengthening the Reporting of Observational Studies in Epidemiology (STROBE) guidelines. Data were obtained from the 2023 National Survey of Children’s Health (NSCH), conducted by the U.S. Census Bureau and sponsored by the Health Resources and Services Administration’s Maternal and Child Health Bureau.^12^ The NSCH provides nationally representative data on children aged 0 to 17 years, focusing on their physical, emotional, and social well-being, family health, and neighborhood environment. The survey uses a complex, stratified sampling design and provides analytic weights that allow extrapolation to the noninstitutionalized U.S. child population.^12^ Since the data are publicly available and de-identified, the study was considered exempt from review by the University of Texas at Austin Institutional Review Board (IRB ID: STUDY00004942).

### Outcome Variables

The primary outcome of interest was household FI, which was assessed by the food situation in a household variable utilizing the NSCH question: “Which of these statements best describes your household’s ability to afford the food you need during the past 12 months?” Respondents selected from the following options: (1) We could always afford to eat good nutritious meals; (2) We could always afford enough to eat but not always the kinds of food we should eat; (3) Sometimes we could not afford enough to eat; or (4) Often we could not afford enough to eat. Although this item is titled “food insufficiency” in the NSCH questionnaire, it captures both the quantity and quality of food available to the household, aligning closely with the U.S. Department of Agriculture’s conceptual definition of FI and the AAP-developed screening toolkit for FI.^1,5^ Prior research has validated this question as an appropriate proxy measure for household FI, as the first response represents households that are food secure, while the latter three reflect households experiencing varying degrees of FI.^13,14^ Accordingly, responses were dichotomized as food secure (option 1) or food insecure (options 2–4), consistent with previous NSCH-based studies.^13–16^

The secondary outcome was overall child health status, based on caregiver responses to the question, “In general, how would you describe this child’s health?” with options of excellent, very good, good, fair, or poor. Following prior literature on pediatric health and national surveys, responses were dichotomized as excellent/very good/good versus fair/poor to identify children with perceived lower health status. ^17^ This variable was analyzed primarily within the subgroup of children with congenital heart disease (CHD) to evaluate the association between FI and overall health status of children with CHD.

### Primary Exposure

The primary exposure of interest was the presence of congenital heart disease. Caregivers were asked, “Has a doctor or other health care provider ever told you that this child has a heart condition?” and, if so, “Was this child born with a heart condition?” Children whose caregivers responded “yes” to both were classified as having CHD, while all others were categorized as non-CHD.^18^

### Covariates

Demographic characteristics included the child’s age in years, sex (male or female), and race/ethnicity (Hispanic, non-Hispanic White, non-Hispanic Black, or other). Family and household characteristics included the U.S. Census region (West, Midwest, North, or South), the age of the first and second adult in the household in years, parental nativity (all parents born in US, any parent born outside US, or child born in US but parents not listed), and family structure (two parents married, two parents unmarried, single parent, grandparent household, or other). Socioeconomic variables comprised the highest household education (less than high school, high school diploma or GED, some college or associate degree, or college degree and above, employment status with at least one caregiver employed (full-time, part-time, or unemployed/retired/working without pay), and the poverty index ratio, calculated based on household income relative to the federal poverty level (FPL), was categorized as ≥400%, 200–399%, 100–199%, or <100%. Receipt of food assistance benefits in the previous 12 months, including Supplemental Nutrition Assistance Program (SNAP/food stamps), Women, Infants, and Children (WIC), and Supplemental Security Income (SSI), was also included to account for program participation among low-income families. These covariates were selected based on prior studies identifying them as key predictors of FI and poor health outcomes among children with congenital heart disease.^2–4, 7,19^

Descriptive analyses were conducted to summarize sociodemographic and clinical characteristics for children with and without CHD. Unweighted frequencies and weighted percentages were reported in accordance with NSCH survey weighting methods.^12^ Bivariate associations between CHD and FI were examined using a survey-weighted chi-square test. Crude odds were calculated, and multivariable logistic regression models were used to estimate adjusted odds ratios (aORs) with 95% confidence intervals (CIs) for the relationship between CHD and FI, controlling for age (child and parent), location, gender, race/ethnicity, education, parental nativity, family structure, receipt of food stamps, WIC benefits, and SSI, and interaction between employment status and poverty index. A second multivariable model was constructed within the CHD subgroup to examine the association between FI and parent-reported overall health status (fair/poor versus good/very good/excellent, controlling for for age (child and parent), location, gender, race/ethnicity, education, parental nativity, family structure, receipt of food stamps, WIC benefits, and SSI, and interaction between employment status and poverty index. A two-tailed alpha level of 0.05 was used to determine statistical significance. All statistical analyses were conducted using SAS version 9.4 (SAS Institute Inc, Cary, North Carolina, USA).

## Results

A total of 53,477 children younger than 17 years were included in the analysis, of whom 1,233 (2.3%) had CHD. Children with CHD had a slightly lower median age (8.1 years [IQR 3.3–12.6]) compared with those without CHD (8.5 years [IQR 4.0–12.8]; p < 0.001). The median age of the first and second adults in CHD households was marginally younger than in non-CHD households (40.2 vs 40.6 years and 39.8 vs 40.5 years, respectively; both p < 0.001). The sex distribution was comparable between groups (49.9% female among CHD vs 48.8% among non-CHD; p = 0.728). Racial and ethnic composition did not differ significantly; approximately one-half of children with CHD were non-Hispanic White (51.5%), followed by Hispanic (26.9%), non-Hispanic Black (10.0%), and other races (11.6%). Geographic distribution varied modestly, with the largest proportion of CHD cases residing in the South (44.9%) and the smallest in the North (14.1%), though this difference was not statistically significant (p = 0.101).

The educational level of the highest-educated adult in the household was similar between groups, with more than half of CHD (57.7%) and non-CHD (54.3%) households reporting a college degree or higher. Family and household structures were comparable: two-parent married households represented 66.6% of CHD families and 66.4% of non-CHD families. Parental nativity did not differ significantly; a slightly larger share of CHD households had all U.S.-born parents (72.5% vs 66.0%; p = 0.140). Economic indicators, including caregiver employment, household income, and public assistance utilization, were largely similar across groups. Participation in food-related assistance programs such as SNAP (17.9% vs 17.3%) and WIC (12.6% vs 10.5%) did not differ by CHD status. However, children with CHD were significantly more likely to receive SSI benefits (5.5% vs 2.4%; p < 0.001). (Table 1)

**Table 1:**
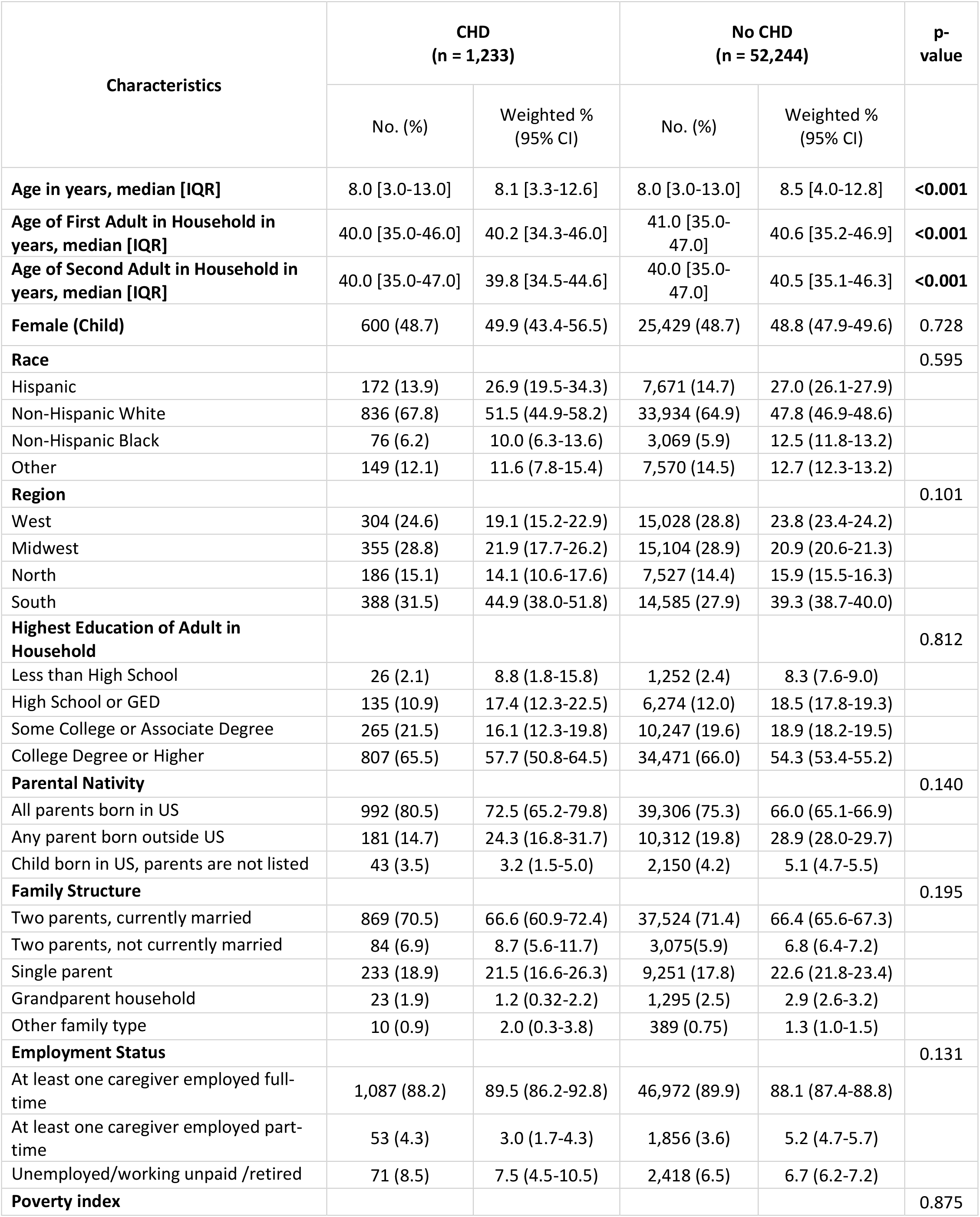
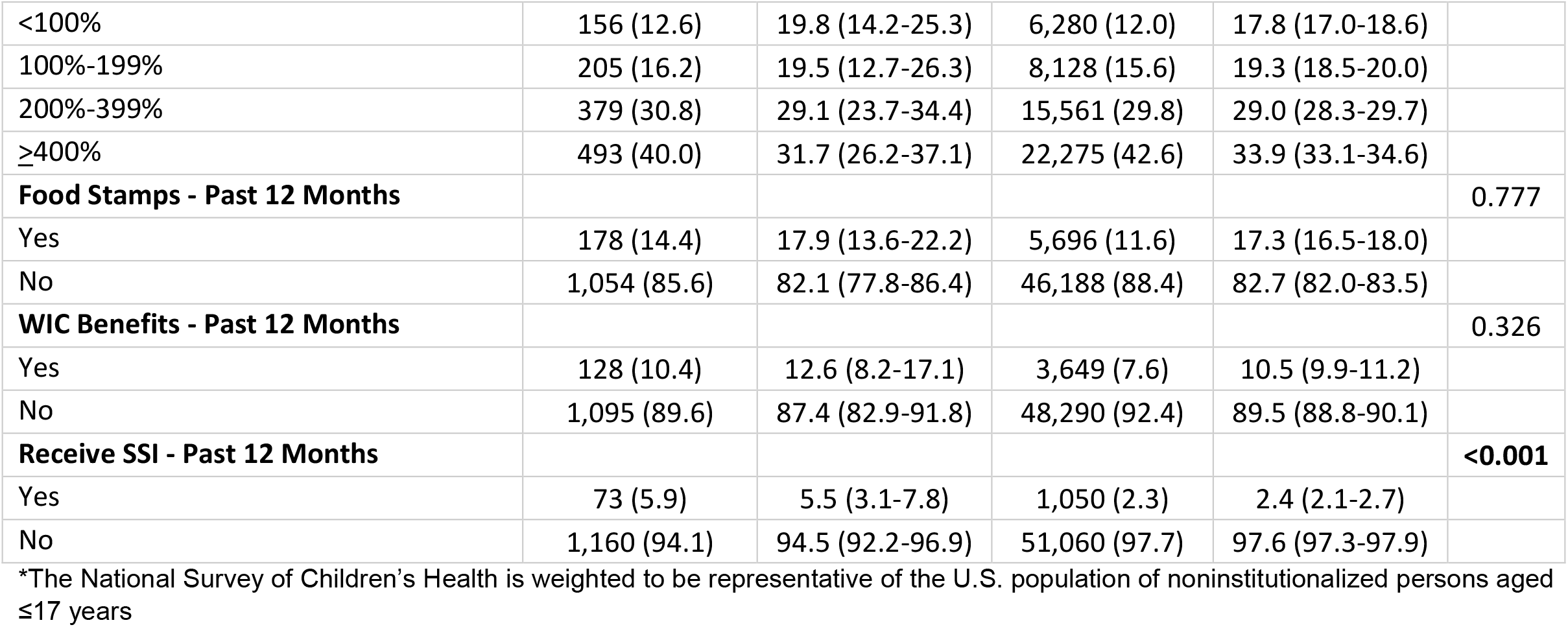
Characteristics of US children with and without congenital heart disease — National Survey of Children’s Health*, United States, 2023.

When examining FI, 40.8% of children with CHD lived in food-insecure households compared with 32.2% of those without CHD (p = 0.005). (Figure 1)

**Figure 1:**
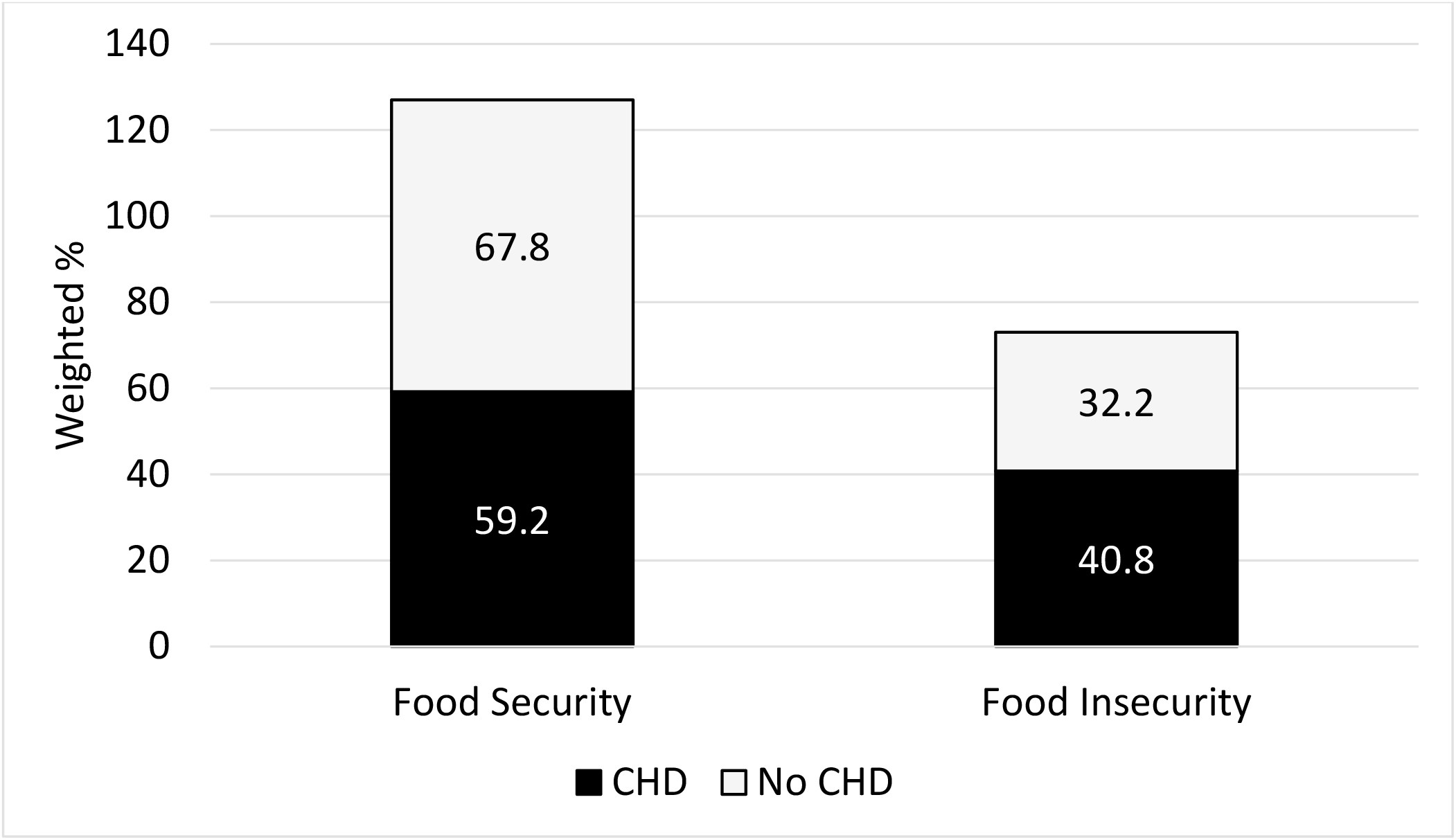
Food Situation by CHD Status— National Survey of Children’s Health*, United States, 2023. *The National Survey of Children’s Health is weighted to be representative of the U.S. population of noninstitutionalized persons aged ≤17 years

In unadjusted analyses, children with CHD had 1.50 times higher odds of experiencing FI (95% CI 1.11–1.80) than children without CHD. After adjustment for demographic, regional, and socioeconomic factors, CHD remained significantly associated with FI (aOR: 1.49, 95% CI: 1.05–2.12; p = 0.027). Several other factors were strongly associated with increased odds of FI: Hispanic ethnicity (aOR: 1.31, 95% CI: 1.14–1.52; p < 0.001), residence in the South (aOR 1.27, 95% CI: 1.20–1.44; p < 0.001), and lower educational level of the household’s highest-educated adult. Compared with households in which an adult held a college degree or higher, those with only a high-school diploma or GED had twice the odds of FI (aOR: 2.01, 95%: CI 1.72–2.33; p < 0.001), and those with some college or an associate degree had similarly elevated odds (aOR: 2.06, 95% CI: 1.83–2.32; p < 0.001). Families living below 200% of the federal poverty level were at particularly high risk, with odds of FI ranging from aOR: 2.75 (95% CI: 1.64–4.62; p < 0.001) for those <100% FPL to aOR: 4.18 (95% CI: 3.02–5.80; p < 0.001) for those between 100–199% FPL. Receipt of public assistance benefits was also associated with higher odds of FI, including SNAP (aOR: 1.74, 95% CI: 1.47–2.07; p < 0.001) and WIC (aOR: 1.23, 95% CI: 1.02–1.48; p =0.027). Conversely, living in a grandparent household (aOR: 0.22, 95% CI: 0.07–0.71; p = 0.011) and having an older adult as the first household member (aOR: 0.98, 95% CI: 0.97–0.99; p < 0.001) were associated with lower odds of FI. (Table2)

**Table 2:**
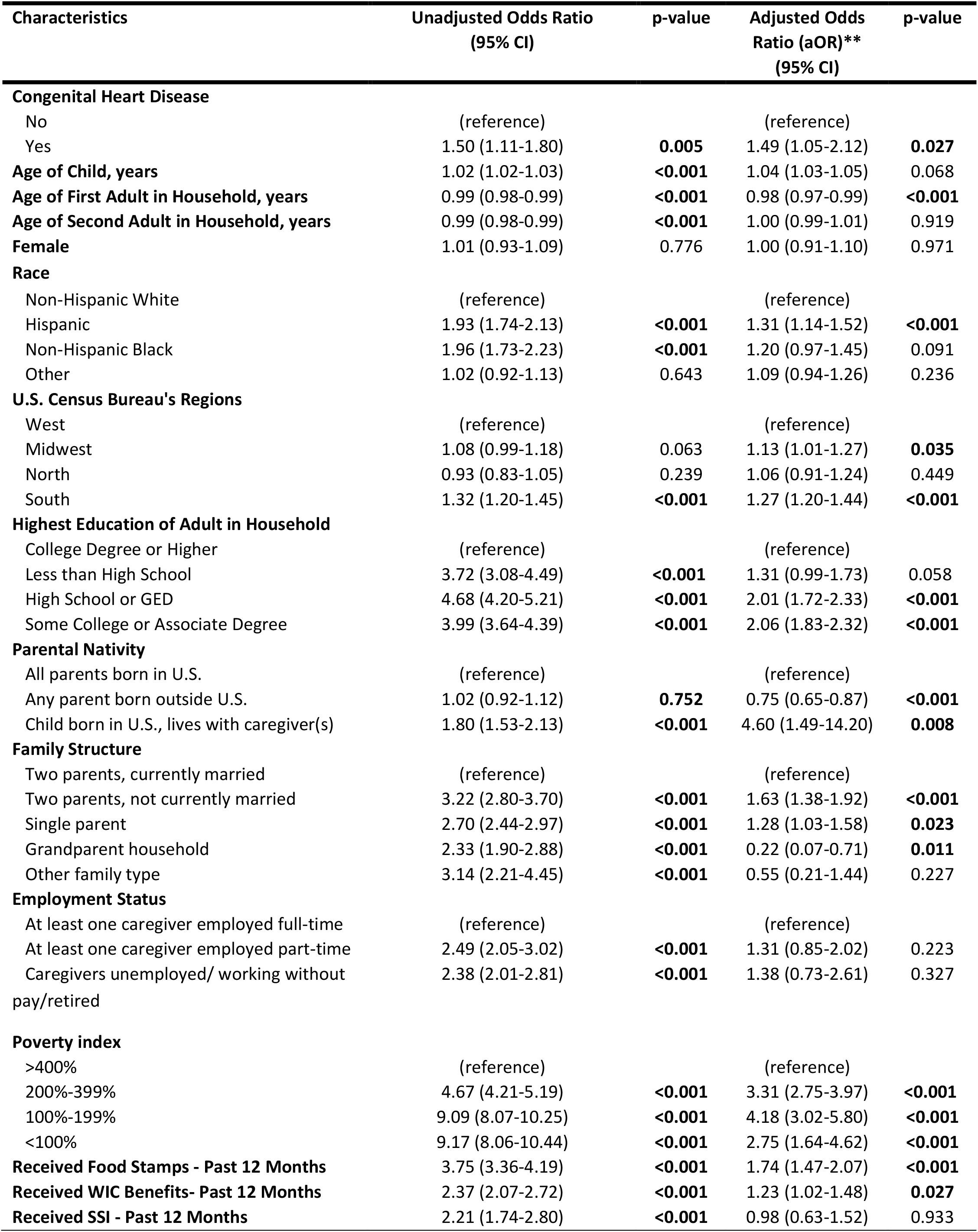
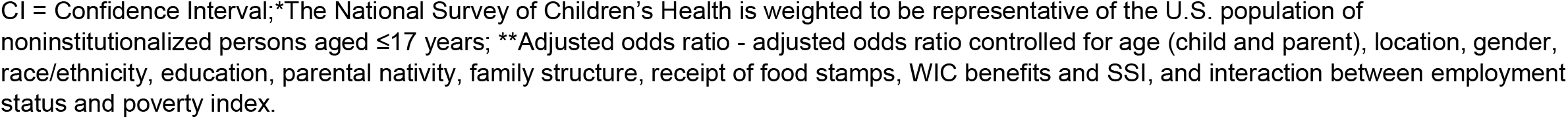
Factors associated with food insecurity— National Survey of Children’s Health*, United States, 2023.

| Characteristics | Unadjusted Odds Ratio<br>(95% CI) | p-value | Adjusted Odds<br>Ratio (aOR)**<br>(95% CI) | p-value |
| --- | --- | --- | --- | --- |
| **Congenital Heart Disease** |  |  |  |  |
| No | (reference) |  | (reference) |  |
| Yes | 1.50 (1.11-1.80) | **0.005** | 1.49 (1.05-2.12) | **0.027** |
| **Age of Child, years** | 1.02 (1.02-1.03) | **<0.001** | 1.04 (1.03-1.05) | 0.068 |
| **Age of First Adult in Household, years** | 0.99 (0.98-0.99) | **<0.001** | 0.98 (0.97-0.99) | **<0.001** |
| **Age of Second Adult in Household, years** | 0.99 (0.98-0.99) | **<0.001** | 1.00 (0.99-1.01) | 0.919 |
| **Female** | 1.01 (0.93-1.09) | 0.776 | 1.00 (0.91-1.10) | 0.971 |
| **Race** |  |  |  |  |
| Non-Hispanic White | (reference) |  | (reference) |  |
| Hispanic | 1.93 (1.74-2.13) | **<0.001** | 1.31 (1.14-1.52) | **<0.001** |
| Non-Hispanic Black | 1.96 (1.73-2.23) | **<0.001** | 1.20 (0.97-1.45) | 0.091 |
| Other | 1.02 (0.92-1.13) | 0.643 | 1.09 (0.94-1.26) | 0.236 |
| **U.S. Census Bureau's Regions** |  |  |  |  |
| West | (reference) |  | (reference) |  |
| Midwest | 1.08 (0.99-1.18) | 0.063 | 1.13 (1.01-1.27) | **0.035** |
| North | 0.93 (0.83-1.05) | 0.239 | 1.06 (0.91-1.24) | 0.449 |
| South | 1.32 (1.20-1.45) | **<0.001** | 1.27 (1.20-1.44) | **<0.001** |
| **Highest Education of Adult in Household** |  |  |  |  |
| College Degree or Higher | (reference) |  | (reference) |  |
| Less than High School | 3.72 (3.08-4.49) | **<0.001** | 1.31 (0.99-1.73) | 0.058 |
| High School or GED | 4.68 (4.20-5.21) | **<0.001** | 2.01 (1.72-2.33) | **<0.001** |
| Some College or Associate Degree | 3.99 (3.64-4.39) | **<0.001** | 2.06 (1.83-2.32) | **<0.001** |
| **Parental Nativity** |  |  |  |  |
| All parents born in U.S. | (reference) |  | (reference) |  |
| Any parent born outside U.S. | 1.02 (0.92-1.12) | **0.752** | 0.75 (0.65-0.87) | **<0.001** |
| Child born in U.S., lives with caregiver(s) | 1.80 (1.53-2.13) | **<0.001** | 4.60 (1.49-14.20) | **0.008** |
| **Family Structure** |  |  |  |  |
| Two parents, currently married | (reference) |  | (reference) |  |
| Two parents, not currently married | 3.22 (2.80-3.70) | **<0.001** | 1.63 (1.38-1.92) | **<0.001** |
| Single parent | 2.70 (2.44-2.97) | **<0.001** | 1.28 (1.03-1.58) | **0.023** |
| Grandparent household | 2.33 (1.90-2.88) | **<0.001** | 0.22 (0.07-0.71) | **0.011** |
| Other family type | 3.14 (2.21-4.45) | **<0.001** | 0.55 (0.21-1.44) | 0.227 |
| **Employment Status** |  |  |  |  |
| At least one caregiver employed full-time | (reference) |  | (reference) |  |
| At least one caregiver employed part-time | 2.49 (2.05-3.02) | **<0.001** | 1.31 (0.85-2.02) | 0.223 |
| Caregivers unemployed/ working without pay/retired | 2.38 (2.01-2.81) | **<0.001** | 1.38 (0.73-2.61) | 0.327 |
| **Poverty index** |  |  |  |  |
| >400% | (reference) |  | (reference) |  |
| 200%-399% | 4.67 (4.21-5.19) | **<0.001** | 3.31 (2.75-3.97) | **<0.001** |
| 100%-199% | 9.09 (8.07-10.25) | **<0.001** | 4.18 (3.02-5.80) | **<0.001** |
| <100% | 9.17 (8.06-10.44) | **<0.001** | 2.75 (1.64-4.62) | **<0.001** |
| **Received Food Stamps - Past 12 Months** | 3.75 (3.36-4.19) | **<0.001** | 1.74 (1.47-2.07) | **<0.001** |
| **Received WIC Benefits- Past 12 Months** | 2.37 (2.07-2.72) | **<0.001** | 1.23 (1.02-1.48) | **0.027** |
| **Received SSI - Past 12 Months** | 2.21 (1.74-2.80) | **<0.001** | 0.98 (0.63-1.52) | 0.933 |

CI = Confidence Interval; \*The National Survey of Children's Health is weighted to be representative of the U.S. population of noninstitutionalized persons aged $\leq 17$ years; \*\*Adjusted odds ratio - adjusted odds ratio controlled for age (child and parent), location, gender, race/ethnicity, education, parental nativity, family structure, receipt of food stamps, WIC benefits and SSI, and interaction between employment status and poverty index.

In the subgroup analysis of children with CHD, 67 (5%) were reported by caregivers to be in fair or poor health. After adjustment for socioeconomic and demographic variables, children with CHD living in food-insecure households had nearly fourfold higher odds of being reported in fair or poor health compared with those in food-secure households (aOR: 3.91, 95% CI: 1.70–9.02; p = 0.001). (Figure 2)

**Figure 2:**
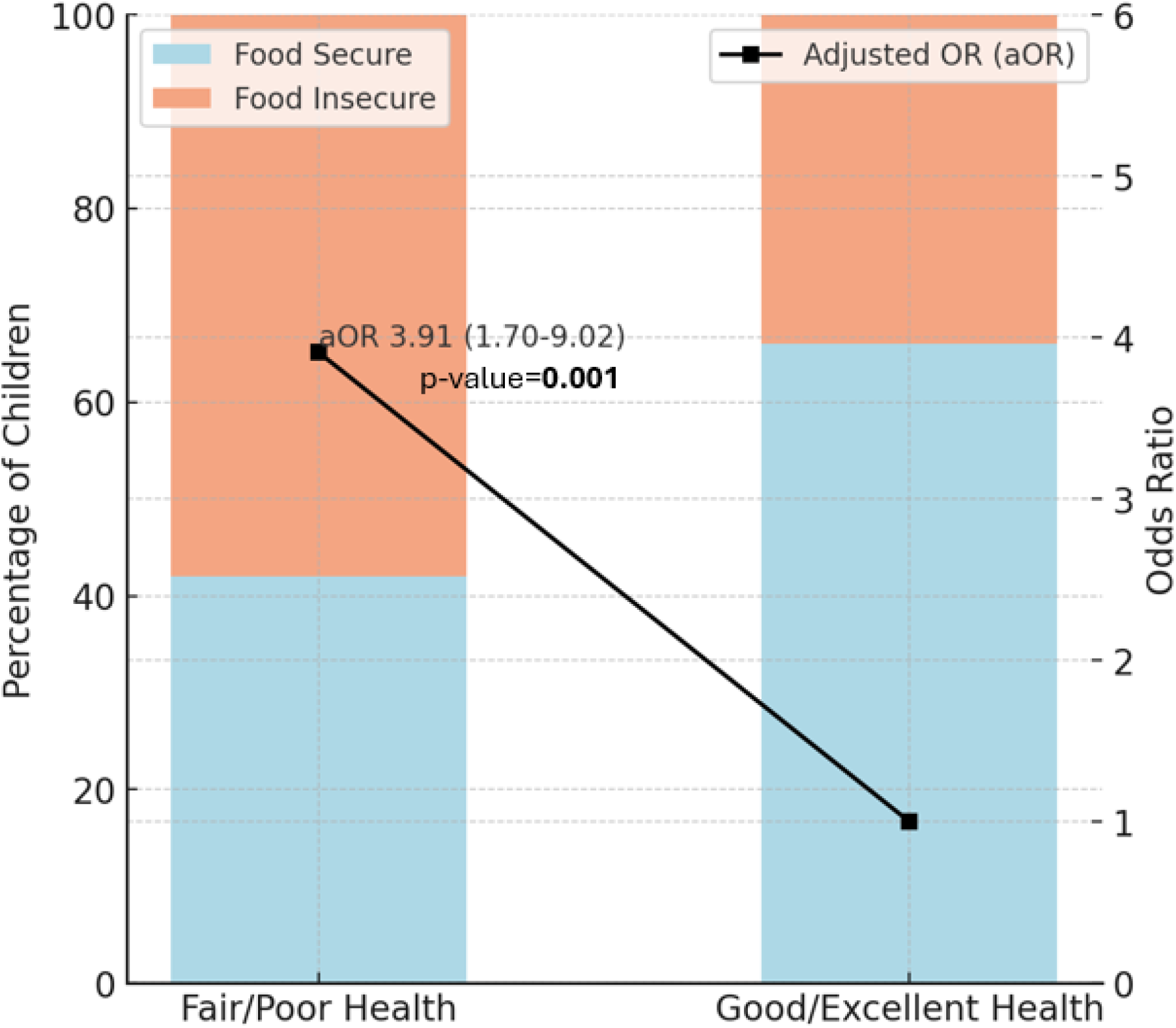
Subgroup Analysis of Overall Health Status of Patients with Congenital Heart Disease— National Survey of Children’s Health*, United States, 2023. ;*The National Survey of Children’s Health is weighted to be representative of the U.S. population of noninstitutionalized persons aged ≤17 years; **Adjusted odds ratio - adjusted odds ratio controlled for age (child and parent), location, gender, race/ethnicity, education, parental nativity, family structure, receipt of food stamps, WIC benefits and SSI, and interaction between employment status and poverty index.

## Discussion

In this nationally representative analysis of U.S. children, those with CHD experienced significantly higher rates of household FI compared to children without CHD, even after adjusting for socioeconomic and demographic factors. Nearly two in five children with CHD lived in food-insecure households, representing a disproportionate burden compared with their healthy peers. Importantly, within the CHD population, FI was independently associated with worse parent-reported overall health status, with affected children nearly four times more likely to be described as being in fair or poor health. These findings highlight the layered vulnerability faced by children with CHD, the chronic medical complexity compounded by adverse social conditions such as FI, which can exacerbate morbidity, and hinder recovery after complex heart procedures.

The relationship between FI and adverse pediatric outcomes has been well documented. FI affects approximately 17% (6.5 million) of U.S. households with children,^1^ and even marginal levels are associated with increased hospitalization, impaired growth, developmental delays, and behavioral and emotional difficulties.^2–4^ Among children with chronic medical conditions, FI amplifies disease-related stressors and increases the likelihood of missed appointments, medication nonadherence, and inadequate nutritional intake.^11^ For children with CHD, whose caloric and micronutrient demands are different from other children of the same age due to altered cardiac physiology,^7,8^ the consequences of FI may be particularly harmful. Previous research has demonstrated that poor nutrition and underweight status are associated with worse surgical outcomes and higher mortality in children with CHD.^20,21^ The current findings extend this literature by demonstrating, at the population level, that FI among children with CHD is not only prevalent but also linked to poorer perceived overall health.

Our results also highlight the strong influence of demographic and socioeconomic factors on FI risk. We found that Hispanic children had 31% higher adjusted odds of FI compared to non-Hispanic White children. Although Black children also experienced higher crude odds, the association attenuated after adjustment. These patterns are consistent with national surveillance showing persistent racial and ethnic disparities in FI, with Hispanic and non-Hispanic Black households facing the highest risk.^1^ Such disparities are rooted in structural racism, discrimination in employment and housing, and unequal access to nutrition programs.^22^ To mitigate these inequities, pediatric cardiology programs should integrate culturally tailored social care interventions such as bilingual FI screening, direct enrollment support for SNAP/WIC, and partnerships with community organizations serving minority populations.

Consistent with national data,^23^ we observed that lower caregiver education and lower household income were disproportionately affected: those with a high school diploma or less had more than twice the odds of FI, and families with incomes below 200% of the federal poverty level were up to four times more likely to experience FI. Regional differences were also prominent with FI being higher in the South and Midwest compared with the West. These disparities likely reflect broader systemic issues such as rural food deserts, limited transportation access, and regional variation in social assistance programs. Addressing these structural inequities will require policy-level interventions that ensure equitable access to affordable, nutritious foods and community-based supports across regions.

Interestingly, we observed that households receiving food stamps or WIC benefits remained at elevated risk of FI, with aORs of 1.74 and 1.23, respectively. This finding aligns with prior studies suggesting that while public nutrition assistance reduces food hardship, it may not be sufficient to fully alleviate FI among medically complex families.^24, 25^ For families caring for children with CHD, fixed benefits may not account for increased expenses related to medical care, transportation, or specialized diets. Clinicians should therefore interpret FI screening results within the broader financial context of the household and advocate for programmatic adjustments that account for disease-related costs. Expanding eligibility thresholds and increasing benefit amounts for families of children with chronic illnesses could meaningfully improve food security and health outcomes.

In our subgroup analysis, 5% of children with CHD reported fair or poor overall health, and those experiencing FI were nearly four times more likely to report poor health. Studies among general pediatric populations have demonstrated that even marginal FI correlates with lower health-related quality of life, increased chronic disease burden, and poorer psychosocial outcomes. ^3,4,10,11^ Especially among children with CHD whose growth, recovery, and neurodevelopment are already fragile, these effects of FI may be disastrous for their recovery. Nutritional inadequacy can compromise wound healing and cardiac function, while caregiver stress and financial strain may impair adherence to complex treatment regimens. Thus, FI likely represents both a marker and a driver of poor health outcomes in the CHD population.

Given these findings, routine screening for FI should be standard practice in pediatric cardiology care. Since 2021, the American Academy of Pediatrics and the Food Research and Action Center have recommended universal FI screening using the validated Hunger Vital Sign™ two-item questionnaire.. ^5^ However, implementation in subspecialty clinics remains limited. Cardiology clinics, where families of children with CHD have longitudinal contact, provide an ideal setting for screening and immediate referral to resources such as hospital-based food pantries, community nutrition programs, and financial counseling. Embedding FI screening into the electronic health record, coupled with care coordination support from social workers or community health workers, could ensure that identified families receive timely assistance.

Beyond screening, our findings emphasize the importance of multilevel interventions that address both immediate nutritional needs and broader socioeconomic determinants. Evidence-based strategies include hospital-community partnerships with local food banks, food prescription initiatives offering vouchers for groceries, and philanthropic collaborations providing medically tailored meals for families of children with complex conditions. Policy and community engagement are also critical to sustaining these efforts. Regional public health agencies should collaborate with healthcare systems to address these issues, while philanthropic organizations can play a vital role in supporting family resource funds and emergency food assistance. Legislative actions that expand funding for WIC, SNAP, and school meal programs are particularly important in mitigating the long-term effects of FI in children with chronic health conditions, especially CHD. By fostering cross-sector collaboration and linking cardiology care, public health, and community nutrition systems, we can move toward an integrated model of care that recognizes food security as a foundational determinant of pediatric cardiac health.

This study has several strengths, including the use of nationally representative data, robust adjustment for confounders, and inclusion of both health and socioeconomic outcomes. However, several limitations merit acknowledgment. The NSCH data relies on caregiver self-report, which may introduce recall bias. The cross-sectional nature limits assessment of temporality between FI and health outcomes. Also, CHD severity and dietary quality were not captured. Despite these limitations, the findings highlight a crucial yet underrecognized aspect of pediatric cardiac care, that food insecurity as a modifiable determinant of health equity.

## Conclusion

Our study demonstrates that children with CHD are at markedly higher risk for food insecurity and that FI is strongly associated with poorer overall health. Socioeconomic disadvantages, lower educational attainment, and regional disparities are key drivers of this relationship. Incorporating routine FI screening into pediatric cardiology care, supported by community partnerships and policy-level interventions, is essential to address this critical social determinant. Through coordinated efforts among healthcare providers, social services, and community organizations, we could ensure that all children with CHD not only survive but thrive towards their road to recovery.

## Data Availability

NSCH data is publicly available.

## Abbreviations

FI: Food Insecurity
CHD: Congenital Heart Disease
NSCH: National Survey of Children’s Health
AAP: American Academy of Pediatrics
STROBE: Strengthening the Reporting of Observational Studies in Epidemiology
FPL: Federal Poverty Level
SNAP: Supplemental Nutrition Assistance Program
WIC: Women, Infants, and Children
SSI: Supplemental Security Income
OR: Odds Ratio
aOR: adjusted Odds Ratio
CI: Confidence Interval

